# Diagnostic performance of multifocal photopic negative response, pattern electroretinogram and optical coherence tomography in glaucoma

**DOI:** 10.1101/2020.03.18.20034785

**Authors:** Khaldoon O. Al-Nosairy, Hagen Thieme, Michael B. Hoffmann

**Author notes:** **Correspondent details:** Michael B. Hoffmann, Department of Ophthalmology, Otto-von-Guericke University, Leipziger Str. 44, 39120 Magdeburg, Germany.

## Abstract

**Objectives:** To optimize stimulation parameters for electroretionographic recordings of the multifocal photopic negative response (mfPhNR) for the detection of glaucoma and to compare the diagnostic accuracy of electrophysiological, structural and functional measures of glaucoma.

**Methods:** In 24 healthy controls, 10 glaucoma suspects (GLA_S_) and 16 glaucoma participants (GLA_G_), mfPhNR for 6 different stimulation rates were assessed to compare their discrimination performance. Subsequently, a cross-modal comparison of the mfPhNR/b-wave ratio was performed with pattern electroretinogram (PERG), and peripapillary retinal nerve fiber layer (pRNFL) thickness. These analyses were based on area under curves (AUC) obtained from receiver-operating-characteristics (ROC) analyses and step-wise regression analyses.

**Results:** Compared to the other mfPhNR-conditions, the PhNR/b-wave ratio for the fastest stimulation condition had the highest AUC for GLA_S_(0.84, P = 0.008, 95%CI: 0.71-0.98); the other modalities, i.e., PERG-amplitude and pRNFL had AUCs of 0.77, and 0.74 respectively. pRNFL was the significant predictor for mfPhNR/b-wave ratio [t (48) = 4, P 0.0002].

**Conclusions:** Fast mfPhNR protocols outperform other mf-protocols in the identification of glaucomatous damage especially for GLA_S_ and thus aid the early detection of glaucoma.

**Significance:** mfPhNR recordings might serve as surrogate marker of ganglion cell dysfunction especially in glaucoma suspects.

## 1 INTRODUCTION

Glaucoma is leading as an irreversible cause of visual impairment(Flaxman et al. 2017) and early detection is of great importance. Several methods are at hand to tap functional and structural damage in glaucoma. Standard automated perimetry (SAP) allows for an assessment of visual function in glaucoma that might be linearly correlated with (Harwerth et al. 2004) or preceded(Quigley et al. 1989; Kerrigan-Baumrind et al. 2000) by structural damage in retina. However, the subjective nature and high variability of SAP might hinder its ability to assess changes in visual function(Chauhan et al. 2008). Electrophysiological measure of vision, on the other hand, is another tool that allows for an objective assessment of visual function.

An important tool for the assessment of retinal ganglion cell function is the steady state pattern electroretinogram (ssPERG) and it can detect glaucoma prior to visual field loss(Bach and Hoffmann 2006, 2008). In fact, previous studies demonstrated that PERG could predict glaucoma conversion of ocular hypertension 4 years before conversion, i.e. before any detectable visual field defects on SAP(Bach et al. 2006; Bode et al. 2011). The full-field ERG (ff-ERG) recorded under light adaptation is another method of particular interest as it provides assessment of inner retinal functions (i.e., ganglion cells) via a component termed the photopic negative response (PhNR)(Viswanathan et al. 1999, 2001). It has the additional benefit, that it is, compared to the PERG, more robust to optic media opacities and refractive errors. PhNRs to full field stimulation do not allow for the detection of focal or localized retinal damage, as the response is the sum response of both healthy and diseased retinal areas(Viswanathan et al. 1999).

Focal PhNRs from multiple retinal areas were reported to detect early functional loss in diseases affecting retinal ganglion cells’ functions(Machida et al. 2010). The multifocal stimulation approach introduced by Sutter et al(Sutter and Tran 1992; Sutter 2001) allows obtaining visual field topographies of retinal and cortical function within short time. Therefore, combining PhNR-recordings with the multifocal approach might enhance its scope in identifying retinal ganglion cell dysfunction. In fact, several studies employing PhNRs recorded with adapted multifocal ERG protocols (mfPhNR) were conducted in glaucoma and optic nerve lesions(Kamei and Nagasaka 2010, 2014; Kamei et al. 2011; Rajagopalan et al. 2014; Kaneko et al. 2015; Kato et al. 2015; Tanaka et al. 2020) and regional PhNR-changes were demonstrated. Different multifocal stimulation modes were used in these studies, i.e., fast(Kamei and Nagasaka 2010, 2014; Kamei et al. 2011; Kaneko et al. 2015; Kato et al. 2015; Tanaka et al. 2020) (1 to 9 interleaved frames) or slow stimulation sequences(Rajagopalan et al. 2014; Van Alstine and Viswanathan 2017) (around 30 interleaved frames). It is not clear at present, whether any of these modes are of specific advantage for the detection of ganglion cell damage. This prompted our present investigation. We aimed (1) to determine which of the slow and fast stimulation modes yields highest mfPhNRs, (2) which condition performs best for the differentiation of controls and glaucoma patients, and (3) how it compares to other established diagnostic methods [i.e., PERG and peripapillary retinal fiber layer thickness (pRNFL)]. We found the fastest mfPhNR-protocol to be the superior multifocal protocol for the discrimination of GLA_S_.

## 2 MATERIALS AND METHODS

### 2.1. Participants

We included 50 participants in the study as detailed below. Participants gave their written consent to participate in the study. The procedures followed the tenets of the declaration of Helsinki and the protocol was approved by the ethical committee of the Otto-von-Guericke University of Magdeburg, Germany. Participants underwent complete ophthalmic examinations and subjective and objective refractions were assessed for both near and far visual acuity to determine best corrected visual acuity (BCVA). Exclusion criteria were any eye diseases or surgeries except cataract and glaucoma surgery, incipient cataract that did not decrease BCVA < 0.8(Bach and Mathieu 2004) and refractive error exceeding ± 5 D or astigmatism > ± 2 D.). There was no significant difference between age across groups [ANOVA, F (2, 48) = 2.7, P = .08]. Demographic and clinical characteristics of each group are shown in Table 1. All glaucoma participants were under either medical and/or surgical treatment.

**Table 1.** Overview of participants’ characteristics

|  | Control | GLA<sub>S</sub> | GLA<sub>G</sub> | P value* |
| --- | --- | --- | --- | --- |
|  | Mean ± SE | Mean ± SE | Mean ± SE |  |
| Age [years] | 52.12 ± 2.25 | 61.84 ± 1.79 | 57.90 ± 3.73 | 0.080 |
| BCVA<br>[decimal] | 1.11 ± 0.04 | 1.73 ± 0.70 | 0.99 ± 0.04 | 0.172 |
| VF-MD [dB] | .74 ± 0.19 | 2.12 ± 0.34 | 4.44 ± 1.07 | < 0.001 |
| VF-PSD [dB] | 4.9 ± 0.75 | 9.18 ± 1.22 | 32.31 ± 7.85 | < 0.001 |

\*ANOVA test. **GLA<sub>S</sub>**= Glaucoma suspects; **GLA<sub>G</sub>**= Glaucoma group; BCVA= Best-corrected visual acuity in decimal; VF-MD= visual field mean deviations, VF-PSD= visual field pattern standard deviations.

#### Controls

Twenty-four participants (mean age ± standard error (SE): 52.1 ± 2.3 years) with normal visual acuity (VA≥1.0) were included in the study. Only left eyes were included for the analysis and all met the inclusion criteria. 1 participant had no PERG and OCT measurements.

#### Primary open angle glaucoma suspects’ group (GLA_S_)

Left eyes of 10 patients (mean age ± SE: 61.8 ± 1.8 years) were included in the study. All glaucoma suspects’ eyes in our study had an open anterior chamber angle with normal appearance of optic disc and RNFL and either(Prum et al. 2016) (i) a visual field defect suspicious for glaucoma damage (i.e., as defined for GLA_G_ group below) without any other apparent causes (n=4), or (i) consistently elevated intraocular pressure (IOP) > 21 mmHg (n=6).

#### Glaucoma group (GLA_G_)

Sixteen glaucoma patients (mean age ± SE: 57.90 ± 3.8 years) were included in the study. Ten patients with primary open angle glaucoma, 5 patients with normal tension glaucoma and 1 patient with pigment dispersion glaucoma. Glaucoma severity ranges from preperimetric to severe stages. Eyes with worse glaucoma damage was chosen for the analysis and in one participant, both eyes had different glaucoma damage stages and both were included separately (i.e., 17 eyes were included in the analysis). GLA_G_ patients met the following criteria(Preiser et al. 2013): 1) Local notching of optic disc rim or vertical cup disc ratio ≥ 7; 2) Visual field defect with 3 or more adjacent points of ≥ 5 dB loss or two or more adjacent points of ≥ 10 dB loss detected in Standard static white-on-white perimetry. Preperimetric glaucoma eyes had no visual field defects.

### 2.2. Standard automated perimetry

Standard automated perimetry (dG2; dynamic strategy; Goldmann size III; OCTOPUS® Perimeter 101, Haag-Streit International, Switzerland) was tested to the central 30° of VF.

### 2.3. SD-OCT

Peripapillary retinal nerve fiber layer thickness (pRNFL) from a 3.5 mm circle scan centered on optic disc (12° diameter) with 768 A-scan Spectral domain optical coherence tomography (OCT) was performed with the OCT Spectralis(Heidelberg Engineering, Heidelberg, Germany.

### 2.4. Electrophysiological testing

#### Procedure

Two types of ERG-recordings, mfPhNR and PERG, were applied in separate sessions, for better comparability both at the same viewing distance of 33 cm. Binocular mfPhNR and PERG were recorded using DTL (Dawson, trick Litzkow 1979, Thompson, Drasdo, 1987) electrodes (DTL Electrode ERG, Unimed electrode Supplies, Ltd, UK) placed in the upper margin of lower lid. The reference electrode (10 mm diameter; Golden EEG Cup Electrodes, Natus Manufacturing Limited, Ireland), filled with conductive paste (Ten20, WEAVER and Company, USA), was attached to the temple ipsilateral of the corresponding eye. The ground electrode filled with conductive paste was pasted on the forehead. Reference and ground electrodes were attached after skin cleaning with a cleaning paste (skinPure, NIHON KODEN Corporation, Tokyo, Japan) to reduce the resistance of the skin below 5 kOhm. Pupils were dilated when recording mfPhNR with Tropicamide 0.5% (Mydriaticum Stulln® UD, Pharma Stulln GmbH, Germany) to approximately 7 mm. Corneas were locally anaesthetized with Oxybuprocain hydrochlorid 0.4% (Conjuncain® EDO®, Bausch&Lomb GmbH, Germany). Refractive correction optimized for a viewing distance of 33 cm for both mfPhNR and PERG recordings.

#### mfPhNR stimulation, recording and analysis

The stimulus display covered 48° and comprised 5 visual field locations (Figure 1), a central (0° – 5°) and four adjacent (from 5°-24° eccentricity). The 5 fields of this display were stimulated independently with an m-sequence, a pseudo-random succession of 0 (no flash) and 1 (flash) states. In accordance with previous literature(Van Alstine and Viswanathan 2017), we applied an m-sequence length of 2^9^-1 (511) steps, each step lasted 13.3 ms. This resulted in a total recording ranging from 3 minutes and 23 seconds in the slowest protocol and 1 minute and 1 second in the fastest protocol. The testing protocol was subdivided into 16 segments to allow blinking. Recording segments comprising artifacts, e.g. from eye movements or blinking, were re-recorded. VERIS Science 6.4.9d13 (EDI: Electro-Diagnostic Imaging, Redwood City, CA, USA) was used for stimulus delivery and electrophysiological recordings. For each area in the dartboard, the stimulus consisted of either 1, 2, or 5 bright frames, each occurring on 50% of the frame changes (75 Hz) followed by either 8 or 25 dark frames This resulted in 3 fast conditions, i.e., 8 dark frames, with 1, 2 and 5 bright frames for stimulation (C_Fast 1_, C_Fast 2_ and C_Fast_3_, respectively) and 3 slow conditions, i.e., 25 dark frames, with 1, 2 and 5 bright frames for stimulation (C_Slow_1_, C_Slow_2_ and C_Slow_3_, respectively). The averaged stimulation frequency arising ranged from 1.3-4.2 Hz. Each condition was repeated twice in a counterbalanced sequence of conditions. The luminance of [i] white frames (stimulation), [ii] uniform grey background, and [iii] black frames (no stimulation) were set at 200, 100, and 7 cd/m^2^, respectively, as checked with a calibrated photometer (CS-100A photometer; Konica Minolta Holdings, Inc.; Japan). Stimuli were presented on a monochrome CRT-monitor (MDG403, Philips; P45 phosphor) at a frame rate of 75 Hz, while the measurement was checked on a separate control monitor.

**Figure 1.**
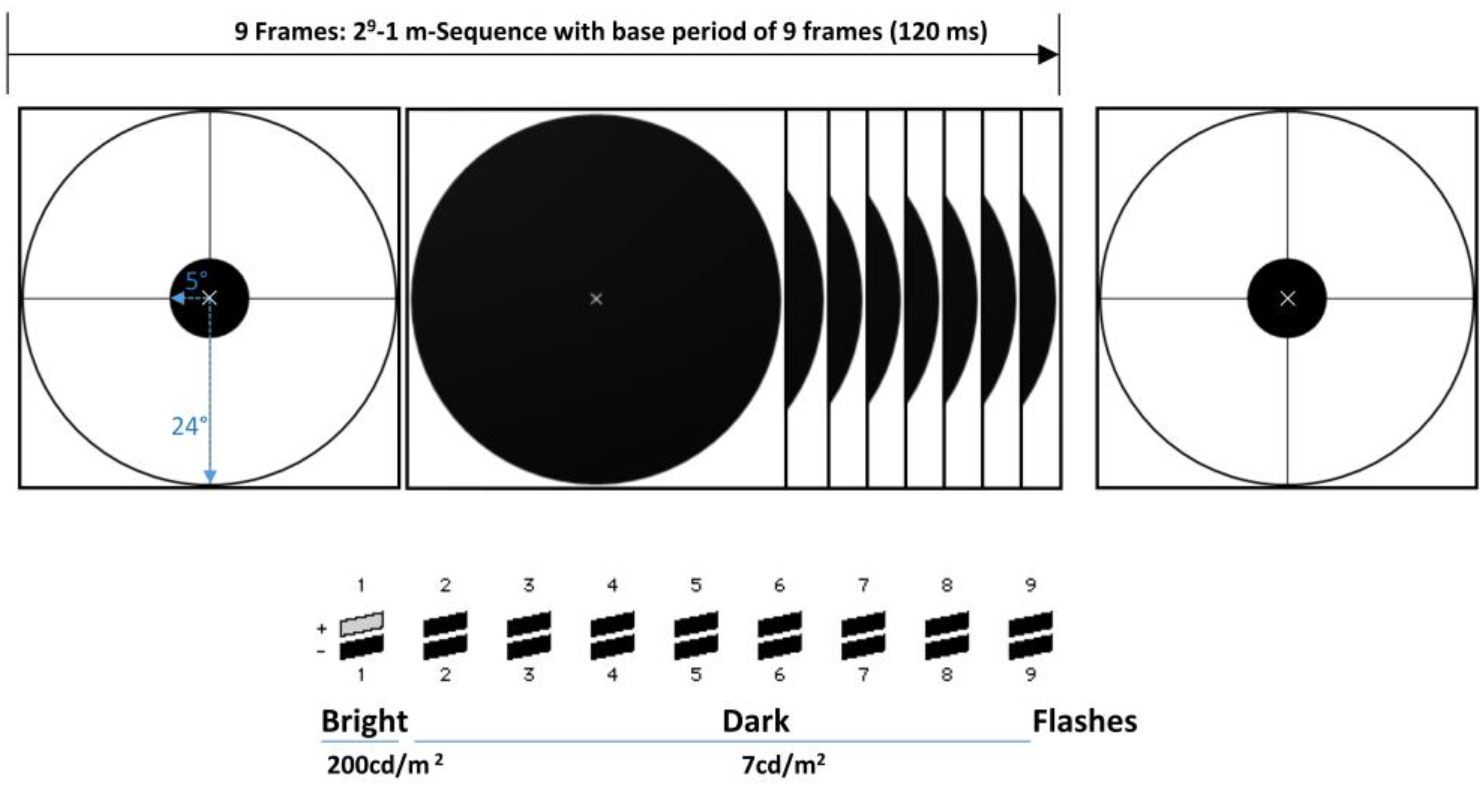
Schematic of the mfPhNR stimulation paradigm. The multifocal stimulus array consisted of 5 visual field locations with inner diameter of 5° and outer of 24° and a small fixation cross in the center. Stimulation followed an m sequence with 2^9^-1 elements, i.e., 511 stimulated patterns of black and white combinations. For the fast sequence protocol (C_Fast_1_) each elements lasted for 9 frames, as depicted.

Using VERIS Science 6.4.9d13, the signals were amplified by 100 K (Grass Model 12, Astro-Med, Inc., West Warwick, RI, USA), band-pass filtered 3-100 Hz and digitized at 1200 Hz. The first order kernels were extracted using VERIS Science 6.4.9d13. Subsequent analyses were performed using Igor (WaveMetrics Inc., Lake Oswego, OR, USA). Traces were then digitally filtered (high pass filter: 3Hz; low pass filter: 45 Hz). Repetitions of each condition were averaged. Traces from right eyes were left-right flipped to match stimulated visual fields of traces recorded from left eyes of other participants.

We determined the amplitudes for the a-wave (1^st^ negativity), b-wave (1^st^ positivity) and the mfPhNR, within a time window of 15 to 35 ms, 20 to 50 ms, and 55 to 90 ms, respectively. For the determination of the latter time window we calculated, following Van Alstine and Viswanathan(Van Alstine and Viswanathan 2017), the waveforms’ grand means for the control group within each age bin and we summed all test location waveforms’ grand means. This resulted in mfPhNRs to peak around 70-75 ms, adding15 ms allowed to cover the individually varying trace forms. Consequently, mfPhNR amplitude was localized within a time window of 55-90, which is similar to conventional PhNR amplitudes timing described previously(Machida et al. 2015). Following studies of mfPhNR(Kaneko et al. 2015) and full field (conventional) photopic ERG(Machida et al. 2008; Preiser et al. 2013), the PhNR amplitude was also evaluated in respect to b-wave measured from baseline (PhNR/b-wave ratio). Before normalizing mfPhNR to the b-wave, we first assessed the b-wave between groups. We found no significant differences of neither b-waves amplitudes nor peak times of all conditions across groups upon one-way ANVOA testing. All amplitudes were measured from trough to baseline. For an assessment of the visual field topography, we performed analyses for different groups of the stimulated visual field locations: We summed all visual field locations to represent the summed visual field locations response (VF_SUM_), summed the pairs of visual field patches to cover four different visual field locations, i.e., upper, lower, nasal and temporal hemifields and periphery), and assessed the central response in isolation.

#### PERG stimulation, recording and analysis

Steady state PERGs were recorded following the international society for clinical electrophysiology of vision (ISCEV) standards for PERG recordings using EP2000 evoked potential system(Bach). By subtending an angle of 62°x49°, the stimulus was presented on 21-inch monochrome monitor with a 75 Hz frame rate (MDG403, Philips; P45 phosphor). Contrast-inverting (15 Hz) checkerboard patterns (mean luminance: 45 cd/m^2^; contrast: 98%) with two checksizes, 0.8° and 15° were presented for stimulation at a viewing distance of 33 cm, as for the mfPhNR measurements. Signals exceeding ± 90 µV were rejected and recollected. Two PERG blocks were recorded and averaged per subject. Only PERG 0.8° amplitude and PERG ratio (small/big checksizes’ amplitudes) were used for analysis. Further description of analysis is mentioned elsewhere(Bach and Hoffmann 2008; Preiser et al. 2013; Al-Nosairy et al. 2020).

### 2.5. Statistics

mfPhNR responses and PERG amplitudes were transferred into Igor sheet and exported to SPSS 26 (statistical Package for the Social Sciences, IBM) or the R statistical system(R Core Team (2013)). Repeated measure analysis of variance (RM-ANOVA), analysis of variance (ANOVA), receiver operating characteristics analysis (ROC) and Post-hoc analysis were run between groups and protocols using SPSS 26 and R. P values were corrected with the Holm Bonferroni correction(Holm 1979) for multiple testing. Pairwise comparisons of two AUCs were conducted to assess whether there is a significant difference by calculating a critical ratio z using the following formula(Hanley and McNeil 1983):

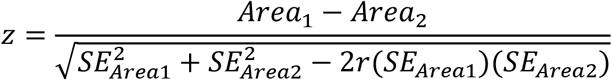

and unpaired comparison of AUCs from 2 different data sets (controls vs GLA_S_ and Controls vs GLA_G_) were done using the following formula(Motulsky):

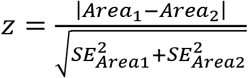

Where A_1_ and A_2_ refers to area under curve with their respective standard error (SE_1_ &SE_2_, respectively) for method 1 and 2, respectively, and r represents estimated correlation between A_1_ and A_2_. Z value is referred to the normal distribution tables and z value ≥ 1.96 means there is a true difference between both AUCs.

## 3 RESULTS

### 3.1. Comparison of mfPhNR recording protocols

Multifocal stimulation settings that were previously applied for mfPhNR recordings ranged between fast (1-9 frames(Kamei and Nagasaka 2010, 2014; Kamei et al. 2011; Kaneko et al. 2015; Kato et al. 2015; Tanaka et al. 2020)) and slow (30 frames(Rajagopalan et al. 2014; Van Alstine and Viswanathan 2017)) sequences. In a first step, we performed a quantitative comparison of these settings and their transitions in order to identify the most effective stimulation condition. This condition subsequently entered the second step, i.e., a detailed analysis of the parameters determining the diagnostic potential.

#### Comparison of stimulation conditions

For an initial characterization of the mfPhNR for the different conditions, we compared response amplitudes between the different conditions for the controls. This was followed by an analysis of the discrimination performance and efficacy in the detection of glaucoma-related damage. We applied six different stimulation settings for mfPhNR recordings and compared the mfPhNR amplitude and mfPhNR/b-wave ratio as detailed in Methods for the control group. A one-way repeated measure ANOVA [factor, *condition* (C_Slow_3_ to C_Fast_1_)] was conducted to determine the effect on mfPhNR amplitude and mfPhNR/b-wave ratio of the summed visual field locations’ response (VF_SUM_). There was a significant effect of condition on both mfPhNR amplitude [F (2.2, 51.7) = 4.3, P = 0.015] and mfPhNR/b-wave ratio [F (1.7, 38.8) = 12.6, P = 0.0001]. Post-hoc analyses were initially performed for the two most widely used conditions (C_Fast_1_ vs C_Slow_1_) and demonstrated both mfPhNR amplitude and mfPhNR/b-wave ratio to be significantly lower for C_Fast_1_ [t (23) = 3.8 and −5.8, P = 0.001 and 0.00002, respectively]. Comparing C_Fast_1_ to the other conditions, mfPhNR-amplitudes were significantly lower than for C_Slow_2_ (P = 0.004), while mfPhNR/b-wave ratios were significantly lower than for all other conditions (see Table 2 for effect sizes). While amplitudes are informative to characterize the responses obtained for different stimulation conditions, an assessment of the discrimination performance between controls vs GLA_S_ and GLA_G_ is instrumental to identify the condition with the highest diagnostic power. Therefore, we conducted ROC analyses and used the area under curve (AUC) as a measure of the diagnostic performance for each condition for both mfPhNR amplitude and mfPhNR/b-wave ratio as depicted in Figure 2. For most conditions, the ROC analyses yielded similar AUCs. Pairwise AUC-comparisons (see Methods), identified only the C_Fast_1_ for the mfPhNR/b-wave ratio to be significantly higher than the C_Slow_1_ in discriminating controls vs GLA_S,_ i.e. AUC ± SE: 0.84 ± .07 vs 0.59 ± 0.1, respectively (P = 0.02). In summary, C_Fast_1_ had the highest discrimination performance, albeit not having the highest amplitudes and therefore entered the subsequent detailed analysis.

**Table 2.**
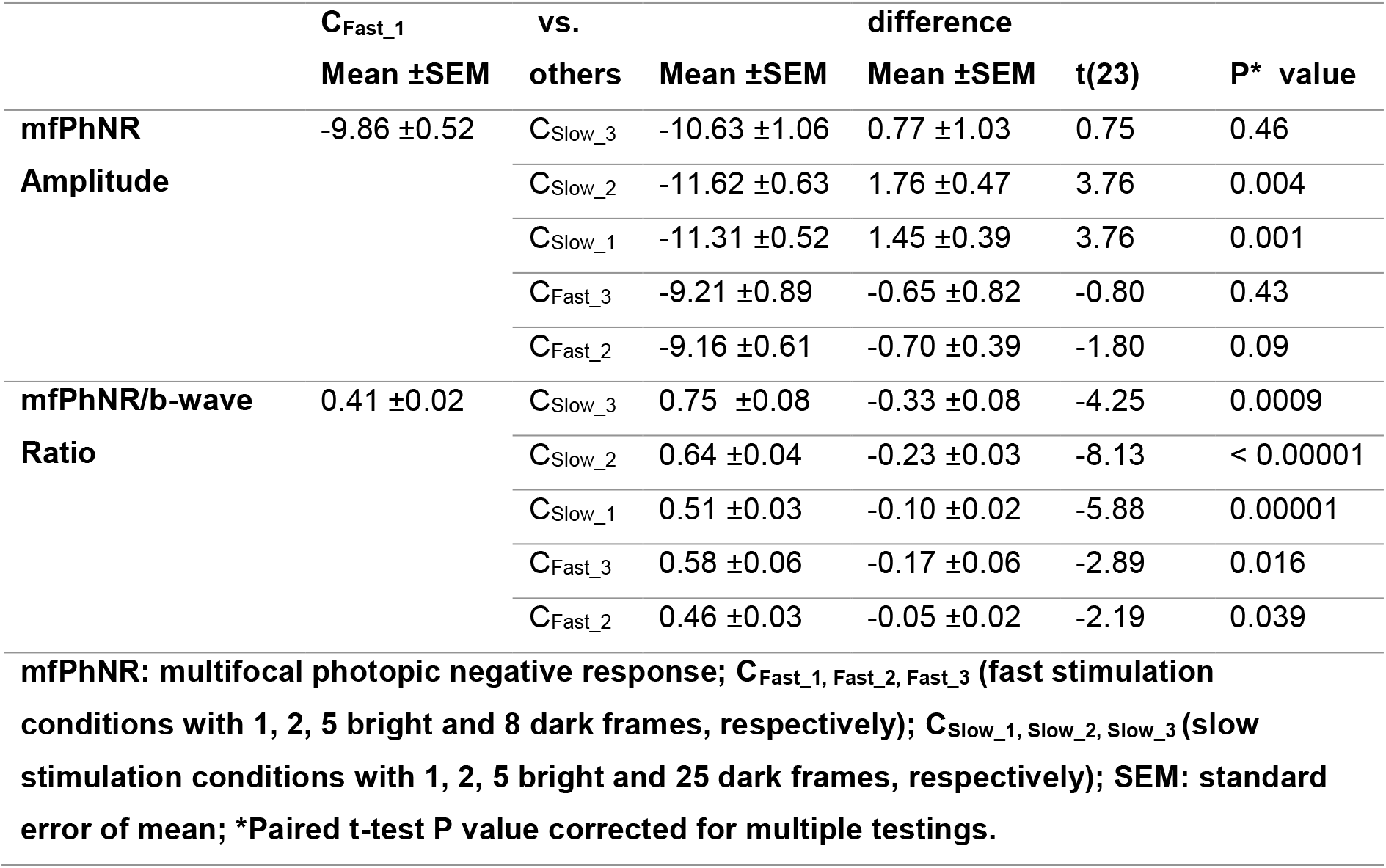
Paired t-test comparisons of mfPhNR amplitudes and ratio of C_Fast_1_vs. other stimulation condition in controls

**Figure 2.**
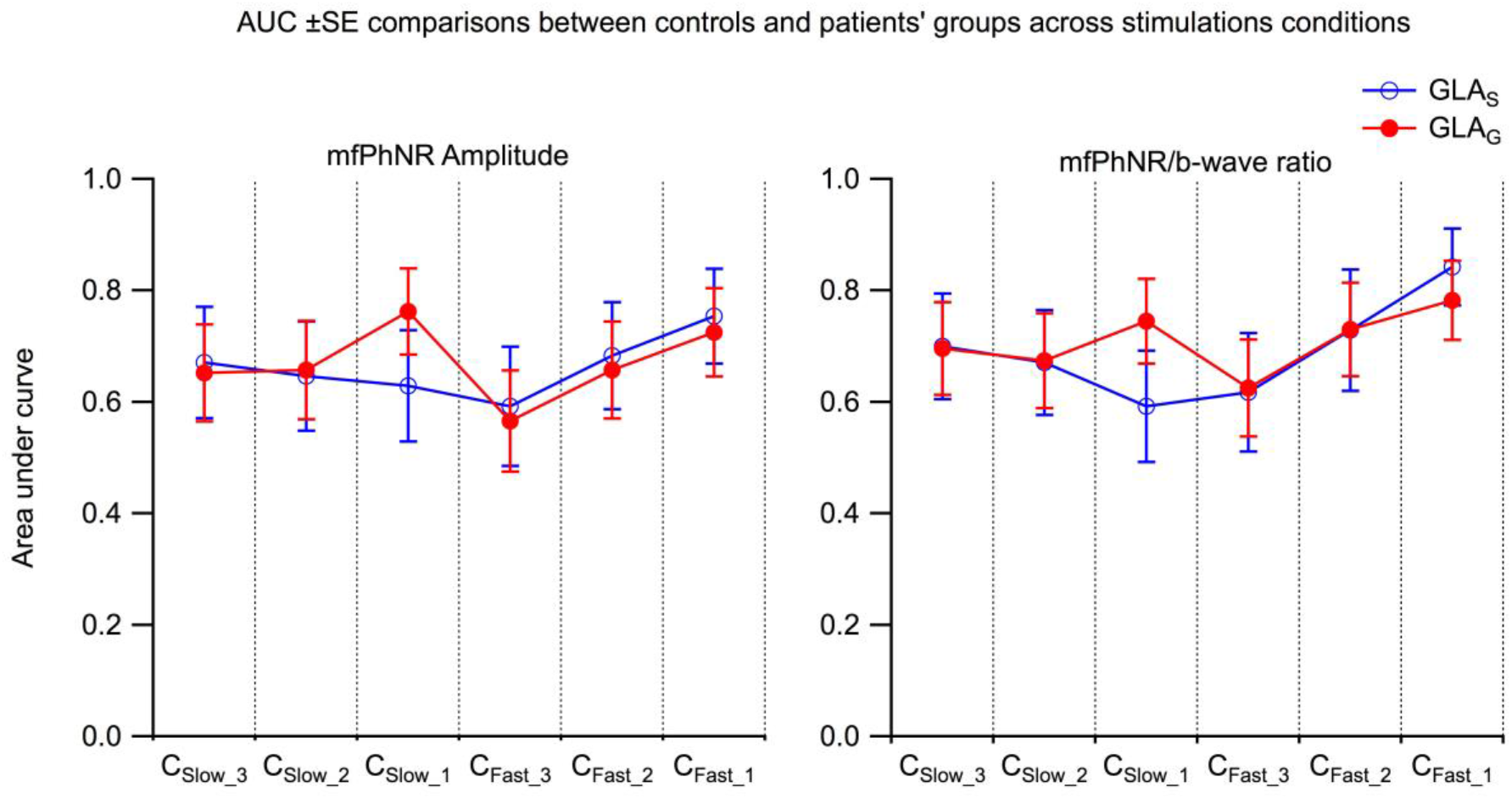
Comparisons of AUCs ± SE for mfPhNR amplitudes (left) and mfPhNR/b-wave ratio (right) for the discrimination between controls vs glaucoma suspects (GLA_S_) and glaucoma (GLA_G_) across a range of stimulations conditions C_Slow_3_ (i.e., slowest condition with 25 dark frames and 5 bright flashes) to C_Fast_1_ (i.e., fastest condition with 8 dark frames and 1 bright flash). While for the mfPhNR amplitudes AUCs did not differ significantly, for the mfPhNR/b-wave ratio AUCs for C_Fast_1_ exceeded those for C_Slow_1_ (P = 0.02).

### 3.2. Trace characteristics and topographical analysis for the C_Fast_1_ protocol

Typical ERG trace shapes were obtained for all participant groups (controls, GLA_S_, and GLA_G_, see Figure 3) with a negativity, positivity and negativity, termed, in accordance with the current literature^21^,a-wave, b-wave, and PhNR, respectively. No significant group effects were observed for peak time and amplitude of a- and b-waves, neither for the responses of each of the five visual field locations, nor for the summed response, i.e., VF_SUM_. In contrast, both the mfPhNR VF_SUM_ amplitude and the mfPhNR/b-wave VF_SUM_ ratio were reduced in the patients groups (GLA_S_ and GLA_G_) compared to controls upon ANOVA testing [mfPhNR VF_SUM_ amplitude: F (2, 48) = 5.7, P= 0.018; mfPhNR/b-wave VF_SUM_ ratio: F (2, 48) = 9.8, P_< 0.007_ = 0.00027]. For subsequent analyses, we focused in accordance to previous studies, on mfPhNR/b-wave ratio of VF_SUM_ response. Finally, no significant group effects were observed for mfPhNR VF_SUM_ peak times [F (2, 48) = 2.9, P ≥ 0.06].

**Figure 3.**
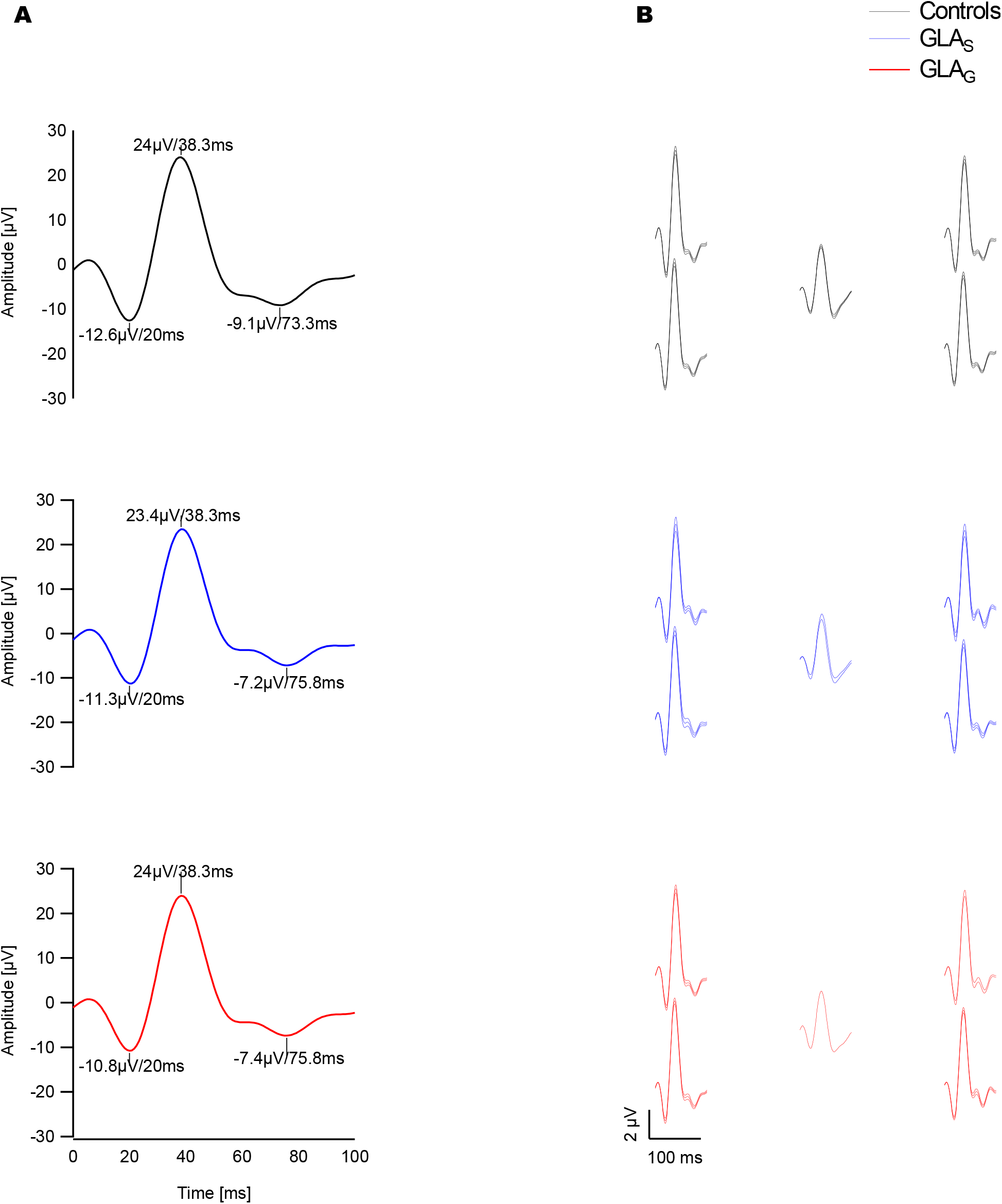
Grand mean average traces of mfPhNR (“VF_SUM_ trace”; A; amplitudes of responses are referenced to the baseline) and trace arrays (B) for controls, glaucoma suspects (GLA_S_) and glaucoma (GLA_G_) of the fastest condition responses (C_Fast_1_).

To assess whether the effects differed for specific hemifields and eccentricities, we subdivided the four peripheral visual field locations into upper, lower, nasal and temporal hemifields and center and periphery. As depicted in Figure 4, in both GLA_S_ and GLA_G_ vs Controls the mfPhNR/b-wave ratio was significantly reduced for all sub-regions analyzed, except the center. The mfPhNR/b wave ratio of the VF_SUM_ response was also significantly different between in both GLA_S_ and GLA_G_ vs Controls upon ANOVA testing [F (2, 48) = 9.8, P_< 0.007_ = 0.00027] as detailed in Figure 4.G.

**Figure 4.**
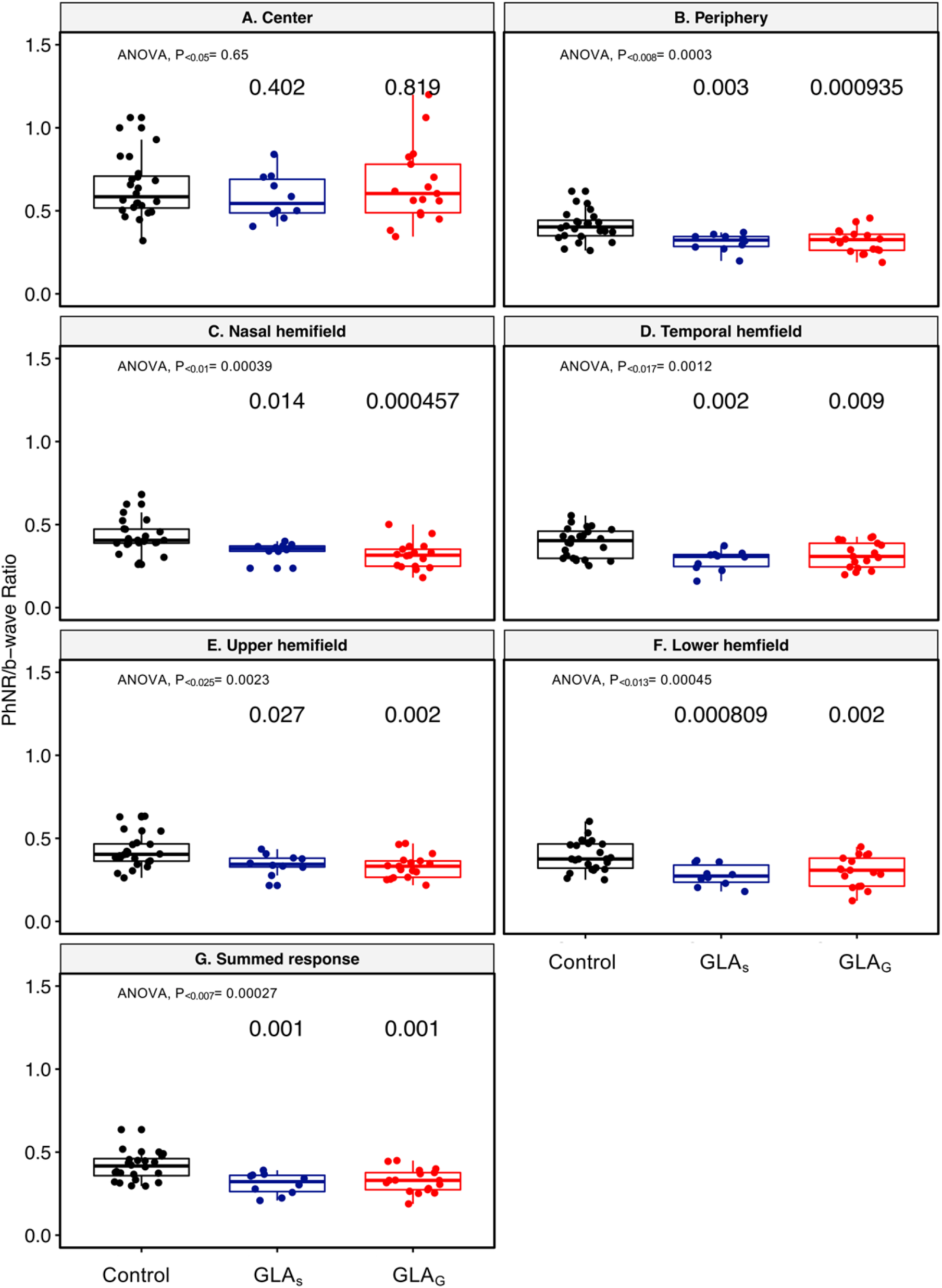
Comparison of mfPhNR/b-wave ratio of the fastest condition across groups and topographic comparisons. Corrected alpha threshold is 0.025 for the more significant P value in each plot. mfPhNR/b-wave ratio is significantly different in all visual field locations across groups except for the center (for P values see panels). Controls’ mfPhNR/b-wave ratio was significantly different vs either group, i.e., glaucoma suspects and glaucoma group. Box: 25%-75% interquartile range; horizontal line is the median value and the whiskers represent the range.

### 3.3. Cross-modal comparison of glaucoma-related damage

We were particularly interested to determine the diagnostic accuracy of our measure, the mfPhNR/b-wave ratio of C_Fast-1_, in comparison to other established diagnostic methods, i.e. PERG or pRNFL. Therefore, we conducted ROC analyses and determined the AUCs (± SE) for mfPhNR/b-wave ratio, PERG .8° amplitude and PERG ratio and pRNFL (Figure 5.A). In GLA_s_, inspection of Figure 5.A suggested that mfPhNR/b-wave ratio was the measure with highest AUC, but statistical testing did not reveal a significant difference between modalities. In GLA_G_, the highest AUC was reached for pRNFL, but without statistically significant differences from other measures. It is noteworthy that all our diagnostic measures showed no AUC difference between Controls vs GLA_S_ and Controls vs GLA_G_ based on unpaired AUC comparison.

**Figure 5.**
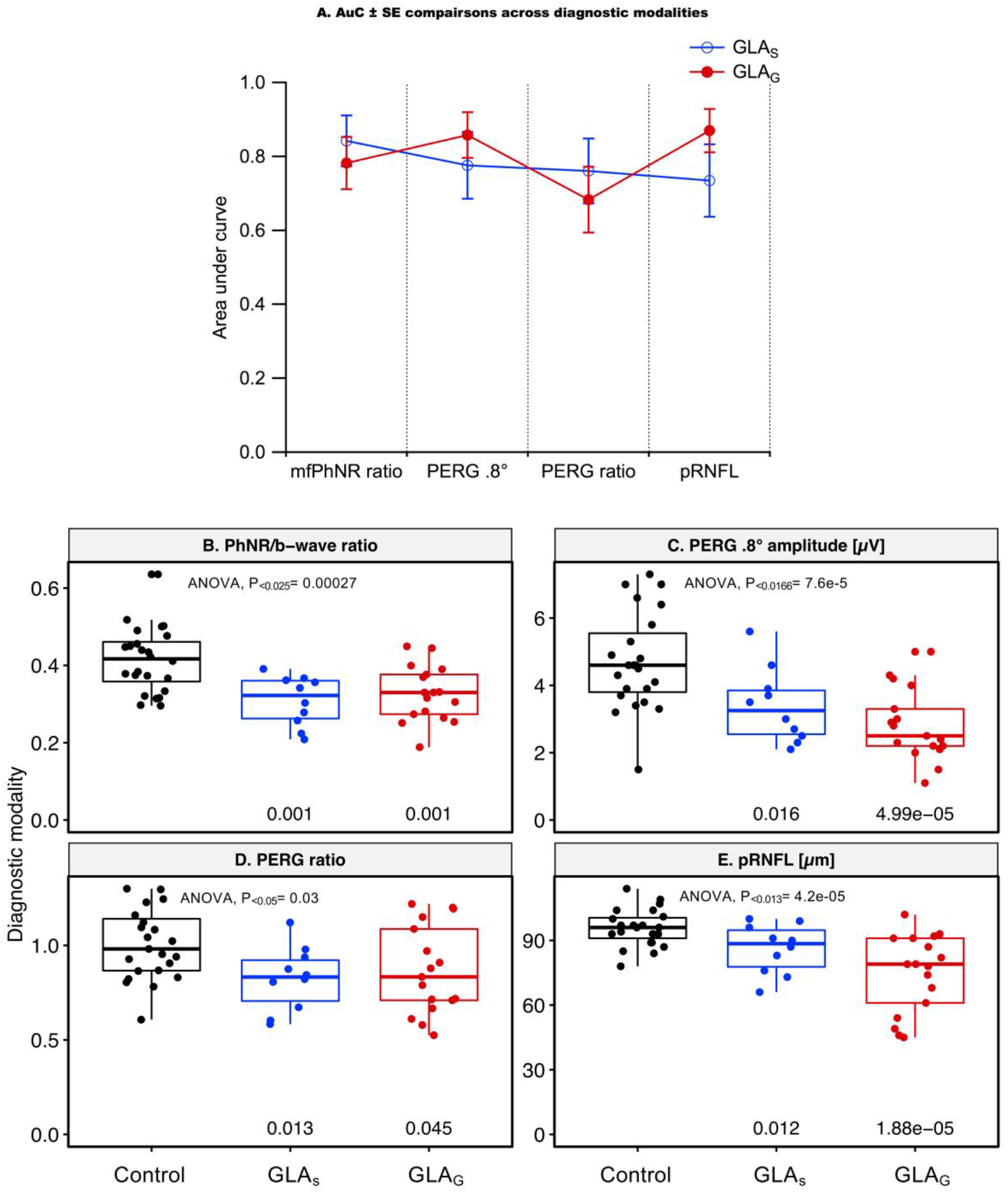
**A** AUC ± SE comparison between diagnostic modalities [i.e., mfPhNR/b-wave ratio of fastest condition C_Fast_1_, pattern electroretinogram (PERG) 0.8° amplitude, PERG ratio and peripapillary retinal nerve fiber layer thickness (pRNFL)] across groups. There were no significant differences of AUCs across modalities after multiple testing correction. Blue= glaucoma suspect (GLA_S_), Red: Glaucoma group (GLA_G_). **B-E** Comparisons of mfPhNR/b-wave ratio, PERG 0.8° amplitude, PERG ratio, and pRNFL across controls, glaucoma suspect and glaucoma group. ANOVA showed significant differences across groups (P values as indicated in the panels). Post-hoc analysis showed significant differences between control vs glaucoma suspect and control vs glaucoma (P values as indicated in the panels). Post hoc analysis corrected alpha value is 0.025 for the more significant P value in each plot. Box: 25%-75% interquartile range; horizontal line is the median value and the whiskers represent the range. Y-axis specified the diagnostic modality as indicated in the panel title.

Subsequently, we compared the functional and structural parameters between both groups applying a one-way ANOVA and post-hoc analyses to compare each patient group vs controls. The mfPhNR/b-wave ratio differed significantly between controls, GLA_S_ and GLA_G_ (P_<0.025_ = 0.00027). Post-hoc tests (Figure 5.B) showed that mfPhNR/b-wave ratio were significantly reduced in GLA_S_ and GLA_G_ vs controls (T-test; P_<0.025_ = 0.00118, P_<0.05_ = 0.00119, respectively). Both PERG 0.8° amplitude and PERG ratio differed also significantly between controls and patient groups (ANOVA; P_<0.0166_ = 0.00008 and P_<0.05_ = 0.03, respectively; Figure 5.C/D). In GLA_S_, PERG 0.8° amplitude and PERG ratio were significantly reduced compared to controls (T-test; P_<0.05_ = 0.015 and P_<0.025_ = 0.013, respectively). In GLA_G_, PERG 0.8° amplitude and PERG ratio were significantly reduced compared to controls (T-test; P_<0.025_ = 0.00005 and P_<0.05_ = 0.045, respectively). Since PERG 0.8° amplitudes had a higher AUC than the PERG-ratio, we used only the PERG 0.8° amplitudes for the below analysis. As a measure of structural damage in glaucoma, mean pRNFL was also statistically different between groups (ANOVA; P_<0.0125_ = 0.000042), it was significantly reduced in GLA_S_ and GLA_G_ vs controls (T-test; P_<0.05_ = 0.012 and P_<0.025_ = 0.00002, respectively; Figure 5.E)

### 3.4. Correlation of PhNR/b-wave ratio and PERG 0.8° amplitude to VF MD, pRNFL thickness, and age

Finally, we tested the correlation of mfPhNR/b-wave ratio with other measures of retinal ganglion cell integrity, i.e. PERG 0.8°, pRNFL and VF-MD. All correlations were significant as detailed in Figure 6. Stepwise linear regression was conducted to identify the measure of ganglion cell integrity predicts the mfPhNR/b-wave ratio best. This analysis indicated pRNFL [t (48) = 4, P = 0.0002] as a single significant predictor for VFSUM mfPhNR/b-wave ratio. Finally, we applied the same analysis steps to PERG 0.8 amplitudes. Significant correlations were found with all other structural and functional measures of retinal ganglion cell integrity, i.e. mfPhNR/b-wave ratio, pRNFL and MD. Again, stepwise linear regression indicated pRNFL [t (48) = 3.4, P = 0.001] as a single significant predictor for PERG 0.8° amplitude.

**Figure 6.**
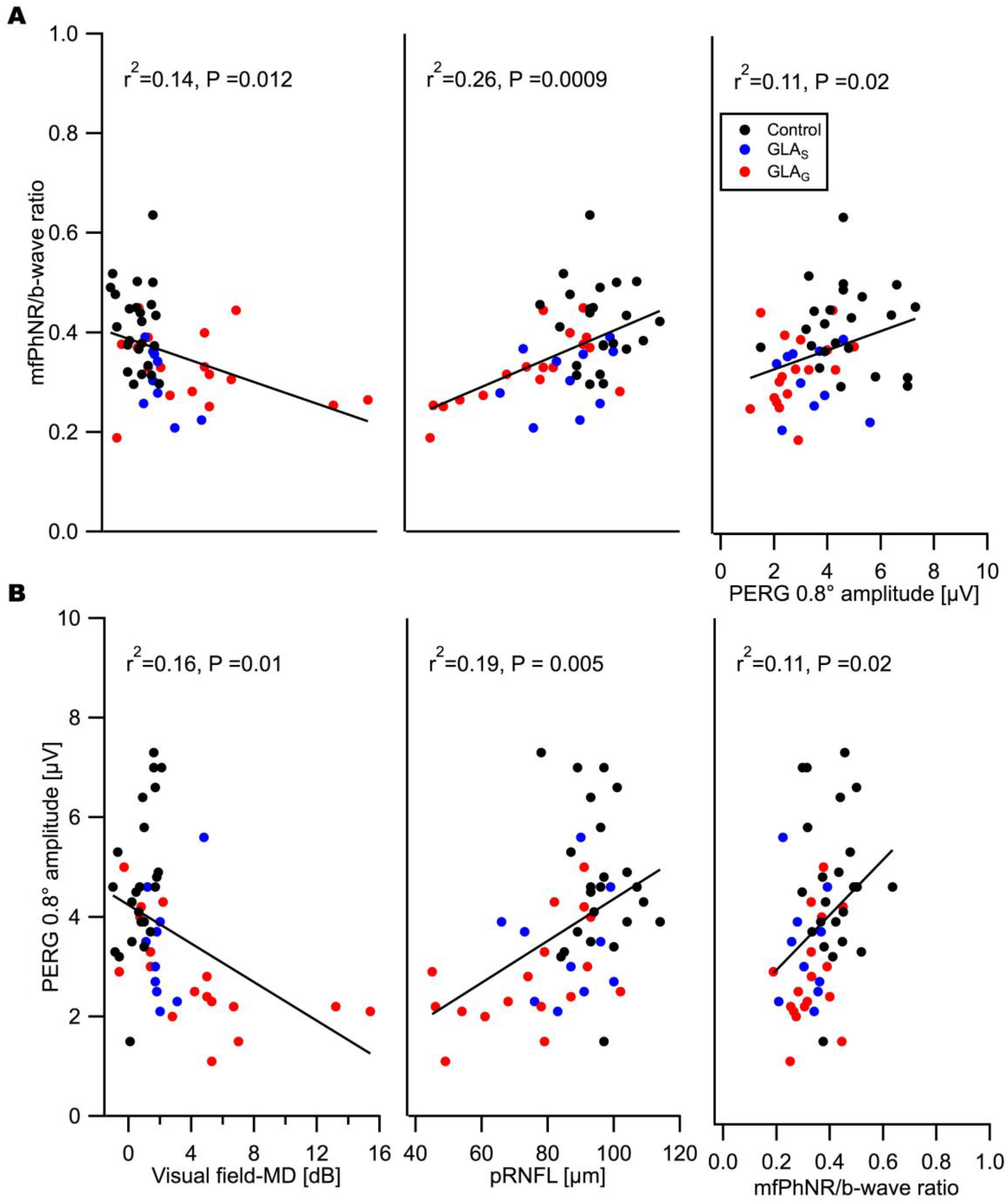
**A** Correlation plots between mfPhNR/b-wave ratio fastest condition (C_Fast_1_) and visual field mean deviation (VF-MD), peripapillary retinal nerve fiber layer thickness (pRNFL) & pattern electroretinogram (PERG) 0.8° amplitude. There were significant association between mfPhNR/b-wave ratio and the above-mentioned measures in which VF-MD, pRNFL and PERG 0.8° amplitude explained 14%, 26% and 11% of mfPhNR/b-wave ratio variance, respectively (see text). **B-**Correlation plots between PERG 0.8° amplitude and VF-MD, pRNFL and mfPhNR/b-wave ratio. There were significant association between PERG 0.8° amplitude and the above-mentioned measures in which VF-MD, pRNFL and mfPhNR/b-wave ratio explained 16%, 19% and 11% PERG 0.8° amplitude, respectively (see text).

## 4 DISCUSSION

### 4.1. Summary of findings

We assessed the effect of stimulus timing on mfPhNR recordings for glaucoma detection and compared a set of conditions with different stimulation rates. Analyzing the mfPhNR/b-wave ratio, we found the best discrimination performance between controls and glaucoma suspects for the fastest stimulation condition, i.e., C_Fast_1_. A spatially resolved analysis of the multifocal responses rendered the analysis of the summed responses of all visual field locations (VF_SUM_) preferable and the central responses least effective. A subsequent detailed assessment of the diagnostic potential of VF_SUM_ mfPhNR/b-wave ratio for C_Fast_1_ revealed a similar potential as PERG, pRNFL and VF-MD to differentiate between Controls and both glaucoma suspects and patients with glaucoma, i.e., GLA_S_ and GLA_G_, respectively.

### 4.2. Comparisons between stimulation conditions

Our comparative approach was motivated by a variety of previous mfPhNR studies on glaucoma detection, that applied using slow (by inserting around 30 frames(Rajagopalan et al. 2014)) or fast sequences (1-9 frames(Kaneko et al. 2015; Kato et al. 2015; Tanaka et al. 2020)). Our study suggests advantages of the faster stimulation protocol (i.e. mfPhNR/b-wave ratio C_Fast_1_) in the early detection of glaucoma, as it performed better than the other conditions tested for the discrimination GLA_S_ from controls. Additionally, it should be noted that C_Fast_1_ is the condition with the shortest recordings time (at equal m-sequence length for all conditions), i.e. 1 minute and 1 second, which bears the potential to increase signal-to-noise and possibly discriminative power further by increasing recording time.

### 4.3. Topographical comparisons of PhNR/b-wave ratio of C_Fast_1_

In the present study, mfPhNR/b-wave ratio of C_Fast_1_ remained unaltered in the central area for both patients’ groups (GLA_S_ and GLA_G_), compared to a decreased response in peripheral visual field locations. Previous studies reported reduced central retinal responses and unaltered peripheral mfPhNR/b-wave ratios glaucoma(Viswanathan et al. 2001; Kaneko et al. 2015; Kato et al. 2015). Recently the mfPhNR/b-wave ratio was reported(Tanaka et al. 2020) to be significantly different in all visual field locations, i.e., both central and peripheral responses in glaucoma patients [range of perimetric damage mean deviation[dB]: −29.26 – 2.02]. This might be related to methodological discrepancies between the studies possibly with diverging sensitivities to different damage sites.

### 4.4. PhNR ratio AUCs for GLA_S_ and GLA_G_

We obtained, based on the mfPhNR/b-wave ratio, similar AUCs for GLA_S_ and GLA_G_ (AUCs: 0.84 and 0.78, respectively). These AUCs correspond well to those of previous studies employing conventional or focal recordings schemes, e.g. Preiser et al (Preiser et al. 2013) (AUC = 0.80), Machida et al(Machida et al. 2010) (AUC = 0.89 to 0.97 for early and late glaucoma, respectively), Kirkiewiez et al. (AUC = 0.78 and 0.86, for moderate and advanced glaucoma, respectively)(Kirkiewicz et al. 2016).

### 4.5. PhNR ratio AUCs comparisons across diagnostic modalities

In our cross-modal comparison, we demonstrated similar AUCs for mfPhNR/b-wave ratio and PERG amplitude (at 0.8° checksize), pRNFL and PERG ratio for GLA_S_ as well as GLA_G_. This is in accordance with Preiser et al.(Preiser et al. 2013) who reported AUCs based on the conventional PhNR/b-wave ratios and the PERG ratio to be comparable for preperimetric (AUCs: 0.80 and 0.73, respectively) and manifest glaucoma (AUCs: 0.80 and 0.79, respectively). By comparison of AUCs of conventional PhNR measures to those of other approaches assessing optic nerve integrity(Kirkiewicz et al. 2016), it was concluded that the conventional PhNR is equivalent to other diagnostic tests in different stages of glaucoma. We consider, therefore, for both conventional and multifocal recording modes, the PhNR/b-wave ratio as a useful measure in glaucoma diagnostics, specifically as it requires no clear media and refraction.

### 4.6. PhNR ratio association with other structural and functional measures

A correlation of PhNR and PERG measures in glaucoma has previously not been reported(Preiser et al. 2013). In our present study, we observed mfPhNR/b-wave ratios to be significantly correlated with both PERG 0.8° amplitude and PERG ratio (r_^2^_ = 0.11, P = 0.02). This supports the view that both measures they reflect similar damage sites and mechanisms, which is further corroborated by the cross-modal correlations of the electrophysiological measures with functional and anatomical measures we observed in accordance with previous conventional PhNR studies(Viswanathan et al. 2001; Machida et al. 2008; Kirkiewicz et al. 2016), PERG studies(Parisi et al. 2001; Toffoli et al. 2002; Elgohary et al. 2019) and another mfPhNR study(Kato et al. 2015). In fact, the observed pRNFL changes were a predictor for both mfPhNR/b-wave ratio and PERG 0.8° amplitude.

## 5. CONCLUSION

We have shown that mfPhNR/b-wave ratio of C_FAST_1_ is complementary in glaucoma diagnosis and found to have equivalent diagnostic accuracy as compared to other established functional and structural methods and might be superior in eyes classified to have normal optic disc (i.e., GLA_S_). These findings open doors to standardizing mfPhNR recording, testing in longitudinal studies with larger sample size and translating them in glaucoma diagnostics.

## Data Availability

Upon request.

## Acknowledgements

This work was supported by European Union’s Horizon 2020 research and innovation programme under the Marie Sklodowska-Curie grant agreement (No. 675033) to Michael B. Hoffmann.

## REFERENCES

Al-Nosairy KO, Bosch JJONV den, Pennisi V, Thieme H, Mansouri K, Choritz L, et al. Interaction of intraocular pressure and ganglion cell function in open angle glaucoma. BioRxiv Prepr. 2020 Jan 31;2020.01.30.924290.

Bach M. Visual Evoked Potentials “EP2000” – Computer system by Michael Bach [Internet]. Available from: https://michaelbach.de/ep2000/

Bach M, Hoffmann M. The origin of the Pattern Electroretinogram. In: Heckenlively JR, Arden GB, editors. Principles and Practice of Clinical Electrophysiology of Vision. 2nd ed. MIT Press; 2006. p. 185–96.

Bach M, Hoffmann MB. Update on the Pattern Electroretinogram in Glaucoma. Optom Vis Sci. 2008 Jun;85(6):386.

Bach M, Mathieu M. Different effect of dioptric defocus vs. light scatter on the Pattern Electroretinogram (PERG). Doc Ophthalmol. 2004 Jan 1;108(1):99–106.

Bach M, Unsoeld AS, Philippin H, Staubach F, Maier P, Walter HS, et al. Pattern ERG as an Early Glaucoma Indicator in Ocular Hypertension: A Long-Term, Prospective Study. Invest Ophthalmol Vis Sci. 2006 Nov 1;47(11):4881–7.

Bode SFN, Jehle T, Bach M. Pattern Electroretinogram in Glaucoma Suspects: New Findings from a Longitudinal Study. Invest Ophthalmol Vis Sci. 2011 Jun 1;52(7):4300–6.

Chauhan BC, Garway-Heath DF, Goñi FJ, Rossetti L, Bengtsson B, Viswanathan AC, et al. Practical recommendations for measuring rates of visual field change in glaucoma. Br J Ophthalmol. 2008 Apr;92(4):569–73.

Elgohary AM, Elbedewy HA, Saad HA, Eid TM. Pattern electroretinogram changes in patients with primary open-angle glaucoma in correlation with visual field and optical coherence tomography changes. Eur J Ophthalmol. 2019 Sep 9;1120672119872606.

Flaxman SR, Bourne RRA, Resnikoff S, Ackland P, Braithwaite T, Cicinelli MV, et al. Global causes of blindness and distance vision impairment 1990–2020: a systematic review and meta-analysis. Lancet Glob Health. 2017 Dec 1;5(12):e1221–34.

Hanley JA, McNeil BJ. A method of comparing the areas under receiver operating characteristic curves derived from the same cases. Radiology. 1983 Sep;148(3):839–43.

Harwerth RS, Carter-Dawson L, Smith EL, Barnes G, Holt WF, Crawford MLJ. Neural Losses Correlated with Visual Losses in Clinical Perimetry. Invest Ophthalmol Vis Sci. 2004 Sep 1;45(9):3152–60.

Holm S. A Simple Sequentially Rejective Multiple Test Procedure. Scand J Stat. 1979;6(2):65–70.

Kamei A, Machida S, Nagasaka E. Multifocal Photopic Negative Response (mfPhNR) and Lineal Visual Sensitivity in Patients with Optic Nerve Lesions. Invest Ophthalmol Vis Sci. 2011 Apr 22;52(14):273–273.

Kamei A, Nagasaka E. Multifocal Photopic Negative Response (mfPhNR) and Retinal Nerve Fiber Layer Thickness (RNFLT) in Patients with Optic Nerve Lesions. Invest Ophthalmol Vis Sci. 2010 Apr 17;51(13):5473–5473.

Kamei A, Nagasaka E. Multifocal Photopic Negative Response (mfPhNR) and Ganglion Cell-Inner Plexiform Layer Thickness (GCIPLT) in Patients with Optic Nerve Lesions. Invest Ophthalmol Vis Sci. 2014 Apr 30;55(13):6236–6236.

Kaneko M, Machida S, Hoshi Y, Kurosaka D. Alterations of Photopic Negative Response of Multifocal Electroretinogram in Patients with Glaucoma. Curr Eye Res. 2015 Jan 2;40(1):77–86.

Kato F, Miura G, Shirato S, Sato E, Yamamoto S. Correlation between N2 amplitude of multifocal ERGs and retinal sensitivity and retinal nerve fiber layer thickness in glaucomatous eyes. Doc Ophthalmol. 2015 Dec;131(3):197–206.

Kerrigan-Baumrind LA, Quigley HA, Pease ME, Kerrigan DF, Mitchell RS. Number of ganglion cells in glaucoma eyes compared with threshold visual field tests in the same persons. Invest Ophthalmol Vis Sci. 2000 Mar;41(3):741–8.

Kirkiewicz M, Lubiński W, Penkala K. Photopic negative response of full-field electroretinography in patients with different stages of glaucomatous optic neuropathy. Doc Ophthalmol. 2016 Feb 1;132(1):57–65.

Machida S, Gotoh Y, Toba Y, Ohtaki A, Kaneko M, Kurosaka D. Correlation between Photopic Negative Response and Retinal Nerve Fiber Layer Thickness and Optic Disc Topography in Glaucomatous Eyes. Investig Opthalmology Vis Sci. 2008 May 1;49(5):2201.

Machida S, Kaneko M, Kurosaka D. Regional Variations in Correlation between Photopic Negative Response of Focal Electoretinograms and Ganglion Cell Complex in Glaucoma. Curr Eye Res. 2015 Apr 3;40(4):439–49.

Machida S, Tamada K, Oikawa T, Yokoyama D, Kaneko M, Kurosaka D. Sensitivity and Specificity of Photopic Negative Response of Focal Electoretinogram to Discriminate Glaucomatous Eyes. Invest Ophthalmol Vis Sci. 2010 Apr 17;51(13):3267–3267.

Motulsky HJ. GraphPad Prism 8 Statistics Guide-Comparing ROC curves [Internet]. [cited 2020 Feb 18]. Available from: https://www.graphpad.com/guides/prism/8/statistics/stat_comparing_roc_curves.htm

Parisi V, Manni G, Centofanti M, Gandolfi SA, Olzi D, Bucci MG. Correlation between optical coherence tomography, pattern electroretinogram, and visual evoked potentials in open-angle glaucoma patients. Ophthalmology. 2001 May;108(5):905–12.

Preiser D, Lagrèze WA, Bach M, Poloschek CM. Photopic negative response versus pattern electroretinogram in early glaucoma. Invest Ophthalmol Vis Sci. 2013 Feb 1;54(2):1182–91.

Prum BE, Lim MC, Mansberger SL, Stein JD, Moroi SE, Gedde SJ, et al. Primary Open-Angle Glaucoma Suspect Preferred Practice Pattern® Guidelines. Ophthalmology. 2016 Jan 1;123(1):P112–51.

Quigley HA, Dunkelberger GR, Green WR. Retinal Ganglion Cell Atrophy Correlated With Automated Perimetry in Human Eyes With Glaucoma. Am J Ophthalmol. 1989 May 1;107(5):453–64.

R Core Team (2013). R: The R Project for Statistical Computing [Internet]. R: A language and environment for statistical computing. R Foundation for Statistical Computing, Vienna, Austria. Available from: https://www.r-project.org/

Rajagopalan L, Patel NB, Viswanathan S, Harwerth RS, Frishman L. Comparison of multifocal photopic negative response (mfPhNR) with structural and functional measures in experimental glaucoma. Invest Ophthalmol Vis Sci. 2014 Apr 30;55(13):5128–5128.

Sutter EE. Imaging visual function with the multifocal m-sequence technique. Vision Res. 2001;41(10–11):1241–55.

Sutter EE, Tran D. The field topography of ERG components in man--I. The photopic luminance response. Vision Res. 1992 Mar;32(3):433–46.

Tanaka H, Ishida K, Ozawa K, Sawada A, Mochizuki K, Yamamoto T. Relationship between structural and functional changes in glaucomatous eyes: A multifocal electroretinogram study [Internet]. In Review; 2020 Jan. Available from: https://www.researchsquare.com/article/c39a0970-ca9d-4cb4-864d-e04c4a860e46/v1

Toffoli G, Vattovani O, Cecchini P, Pastori G, Rinaldi G, Ravalico G. Correlation between the retinal nerve fiber layer thickness and the pattern electroretinogram amplitude. Ophthalmol J Int Ophtalmol Int J Ophthalmol Z Augenheilkd. 2002 Jun;216(3):159–63.

Van Alstine AW, Viswanathan S. Test–retest reliability of the multifocal photopic negative response. Doc Ophthalmol. 2017 Feb;134(1):25–36.

Viswanathan S, Frishman LJ, Robson JG, Harwerth RS, Smith EL. The photopic negative response of the macaque electroretinogram: reduction by experimental glaucoma. Invest Ophthalmol Vis Sci. 1999 May 1;40(6):1124–36.

Viswanathan S, Frishman LJ, Robson JG, Walters JW. The photopic negative response of the flash electroretinogram in primary open angle glaucoma. Invest Ophthalmol Vis Sci. 2001 Feb;42(2):514–22.

